# Identification of lncRNAs differentially expressed in Alzheimer’s disease brains using RNA sequencing

**DOI:** 10.1101/2025.03.31.25324765

**Authors:** M. Muaaz Aslam, Marcel Schilling, Valerija Dobricic, Janina Fuß, Sören Franzenburg, Christina M Lill, Laura Parkkinen, Lars Bertram

## Abstract

Long non-coding RNAs (lncRNAs) have been reported to show differential expression in Alzheimer’s disease (AD), albeit with inconsistent results. In addition, only a few studies explored lncRNA expression in human brain samples. In this study, we performed differential gene expression (DGE) analyses on lncRNA expression data (derived from RNA sequencing [RNA-seq]) from a total of n=1,220 samples collected from three different human brain regions. These included 177 individuals drawn from the OPTIMA study (discovery data; n[AD cases] = 95, n[controls] = 82) collected from entorhinal cortex (EC). In addition, we analyzed independent RNA-seq data from the ROSMAP (dorsolateral prefrontal cortex; n[AD cases] = 436, n[controls] = 333]) and BANNER studies (fusiform gyrus; n[AD cases] = 206, n[controls] = 68). In the discovery dataset (EC), we identified 121 significantly differentially expressed lncRNAs after FDR correction. Meta-analysis with ROSMAP and BANNER data resulted in a total of 356 significantly differentially expressed lncRNAs, showing the same direction of effect in all three datasets. Besides replicating many AD-related lncRNAs, we also identified several dozen lncRNAs not previously linked to AD, including *Lnc-TNFRSF13B-2, CSRP1-AS1, Lnc-MIB2-1, Lnc-TTC5-2, and GSN-AS1.* GO annotation highlighted numerous relevant processes, including neuron development, synapse function, and axon development. To our knowledge, our study comprises the largest RNA-seq-based DGE analysis of the human brain and is the first to utilize EC samples, the site where progressive AD pathology originates. With its large number of novel AD-associated lncRNAs it provides a vastly extended reference for future work on this topic.

## Introduction

Alzheimer’s disease (AD) is a slowly progressing, irreversible, multifactorial, and detrimental neurodegenerative disorder, mainly affecting the elderly. AD is the leading cause of dementia, contributing to roughly 60-80% of all dementia cases^1, 2^. Neuropathologically, the disease is characterized by extracellular amyloid-β (Aβ) plaques, and intracellular neurofibrillary tangles composed of phosphorylated tau protein (p-tau) that, clinically, eventually lead to varied degrees of amnestic and non-amnestic cognitive impairments^3, 4^. AD prevalence doubles approximately every five years after 60 years of age, rising to as much as 40% in those aged 85 and older^5^. The prevalence of AD is much higher in women as compared to men, to the extent that nearly two out of every three AD patients are women^6^. Risk for AD is determined by a complex interaction of genetic, epigenetic, and non-genetic factors with age representing the most significant determinant^7, 8^.

Among the non-genetic factors contributing to AD risk and pathophysiology are long non-coding RNAs (lncRNAs). These molecules represent a class of non-coding RNAs (ncRNAs), which form heterogeneous transcripts of more than two hundred nucleotides in length but with no apparent protein-coding role^9, 10^. The majority of lncRNAs (∼98%) follow a similar general pattern of biogenesis as messenger RNAs (mRNAs), that is, they are transcribed by RNA polymerase II (Pol II), followed by 5’ 7-methyl guanosine capping, 3’ polyadenylation, and splicing through a standard canonical pathway^11, 12^. The stability of lncRNAs varies, with some being stabilized through polyadenylation, while others rely on secondary structures, such as triple-helical formations at their 31 ends^13, 14^. While lncRNAs generally exceed mRNAs in number^15^, they are usually expressed at lower levels than mRNAs^11^. LncRNAs are normally localized to the nucleus (where they unfold their molecular action, see below) and exhibit distinct tissue-specific expression patterns, highlighting their crucial role in tissue-specific processes^11, 12, 16^. Lastly and contrary to mRNAs, which are highly conserved across different species and contain a greater number of exons (typically ≥5), lncRNAs exhibit less sequence conservation and encompass fewer (ranging from 2-3) exons^17–19^.

Mechanistically, lncRNAs play an essential role in regulating gene expression (in *cis* and *trans*), chromatin structure, RNA stability, splicing, and translation through their interaction with DNA, RNA, and proteins^20^. Their multifaceted regulatory capacity makes lncRNAs plausible candidates for contributing to the pathophysiology of various diseases, including AD^21^. To this end, prior work has focused on determining lncRNA expression patterns in AD across various tissue types. For instance, previous studies have reported evidence for differential expression of multiple lncRNAs in AD cases when compared to controls or other disorders, e.g., in plasma^22, 23^, cerebrospinal fluid (CSF)^24^, and human brain samples^25–29^. In the brain, consistent evidence for differential expression was observed with *BACE1-AS*, *HAR1A, LINC01094, LY86-AS1, LINC01007, MIAT, MIR7-3HG, MALAT1* , *MAP4K3-DT, MEG3, MEG8, MEG9, NEAT1, NECTIN3-AS1, STARD4-AS1, PCA3, and XIST* ^25, 30–37^. Furthermore, genome-wide association studies (GWAS) of AD increasingly discover single nucleotide polymorphisms (SNPs) and other types of genetic variants located in non-coding regions of the genome, many mapping to lncRNA loci, further strengthening the notion that this type of RNA plays an important role in AD pathophysiology^19,38, 39^.

While several studies have been published probing for differential expression of lncRNAs in AD vs. controls^22–29^, only a few have utilized RNA sequencing (RNA-seq) in human brain samples^25–29^. Moreover, most of these studies relied on small datasets (n < 100), limiting their ability to provide a comprehensive view of lncRNA (dys)regulation in AD. In this study, we aimed to address and extend this lack of information by applying RNA-seq in post-mortem tissue slices derived from the entorhinal cortex (EC), a brain region typically affected in the early stages of AD, in n = 177 AD patients and controls. These novel lncRNA expression data were meta-analyzed with previously published RNA-seq datasets generated from dorsolateral prefrontal cortex (n = 769; ROSMAP)^40^ and fusiform gyrus (n = 274; BANNER)^41, 42^. Together, we combined molecular data from the brains of n = 1,220 individuals which, to our knowledge, represents the largest RNA-seq-based assessment of lncRNA expression in the field to date. Overall, we identified 356 lncRNAs showing consistent evidence for differential expression in AD across all three datasets, the vast majority not previously implicated in AD pathogenesis. *In silico* functional characterizations using lncRNA-mRNA co-expression and gene ontology (GO) enrichment analysis highlighted synaptic and other neuronal functions as molecular pathways most likely affected by differentially expressed lncRNAs.

## Results

Figure 1 depicts a visual summary of the study design and methods used..

**Figure 1:**
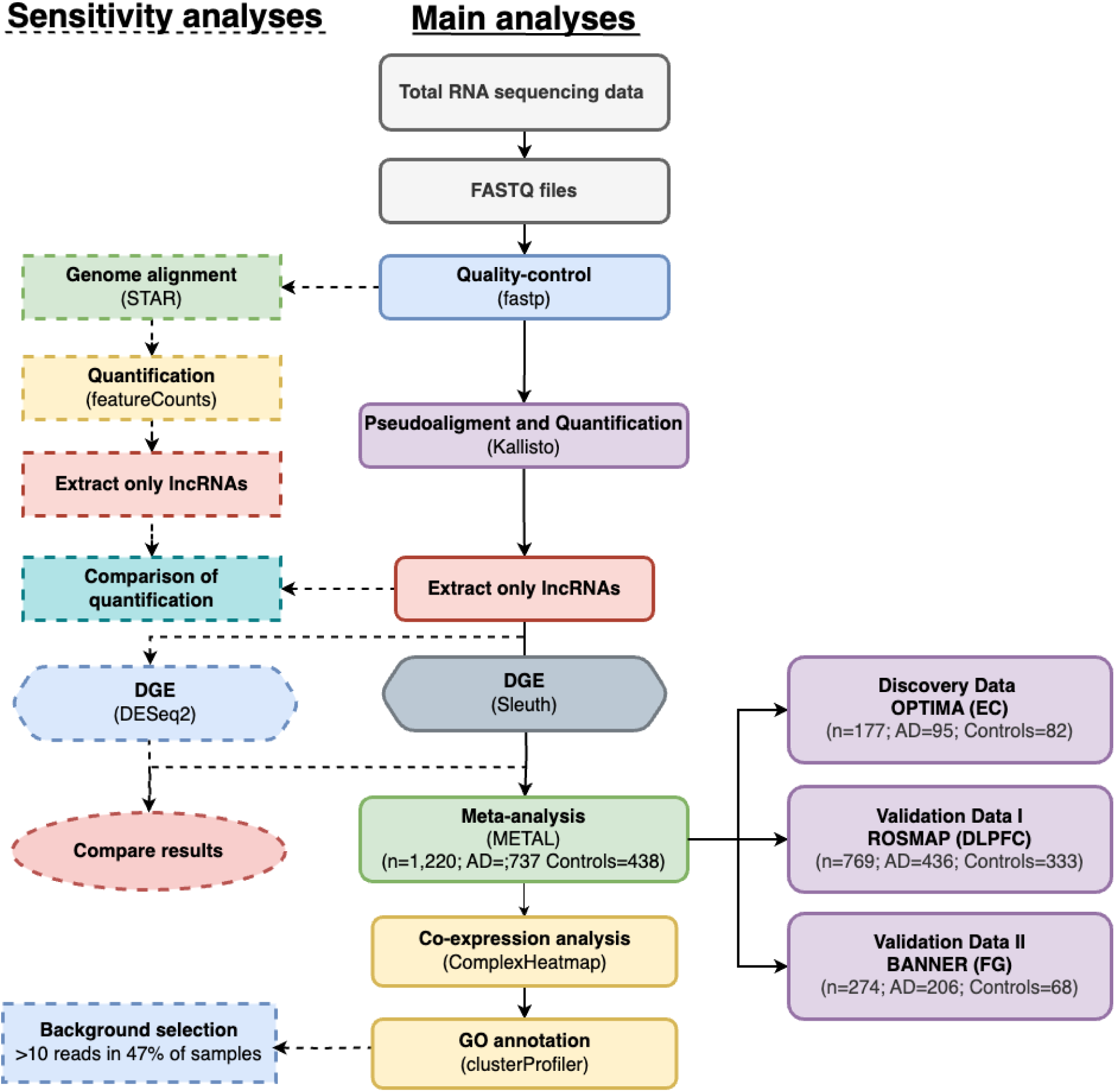
A schematic presentation illustrating the general workflow of this study. Boxes and arrows with broken lines indicate the sensitivity analyses conducted on different data processing steps. EC: Entorhinal cortex; DLPFC: Dorsolateral prefrontal cortex; FG: Fusiform gyrus

### Transcriptome-wide DGE analyses highlight numerous lncRNAs differentially expressed in entorhinal cortex

Demographic details of the 177 OPTIMA study participants analyzed here (n[AD cases] = 95, n[controls] = 82) are shown in **Supplementary Table S1** . Overall, there were no significant differences in the distributions of sex, age at death, RIN, and PMI values between AD cases and controls.

DGE analyses resulted in a total of 1,710 lncRNAs showing nominal significance (p-values [p] ranging from 3.53E-9 to 4.99E-02, λ = 1.18) (**Supplementary Table S2A** ). Of these, the expression of 121 lncRNAs was significantly different in AD vs. controls after FDR (5%) correction (**Supplementary Table S2B, Figure 2** ). Of the 121 FDR-significant lncRNAs, 91 showed a decreased expression in AD vs. controls (q-values [q] ranging from 4.94E-05 to 4.95E-02), while the remaining 30 exhibited evidence for an increased expression (q range = from 4.94E-05 to 4.80E-02).

**Figure 2:**
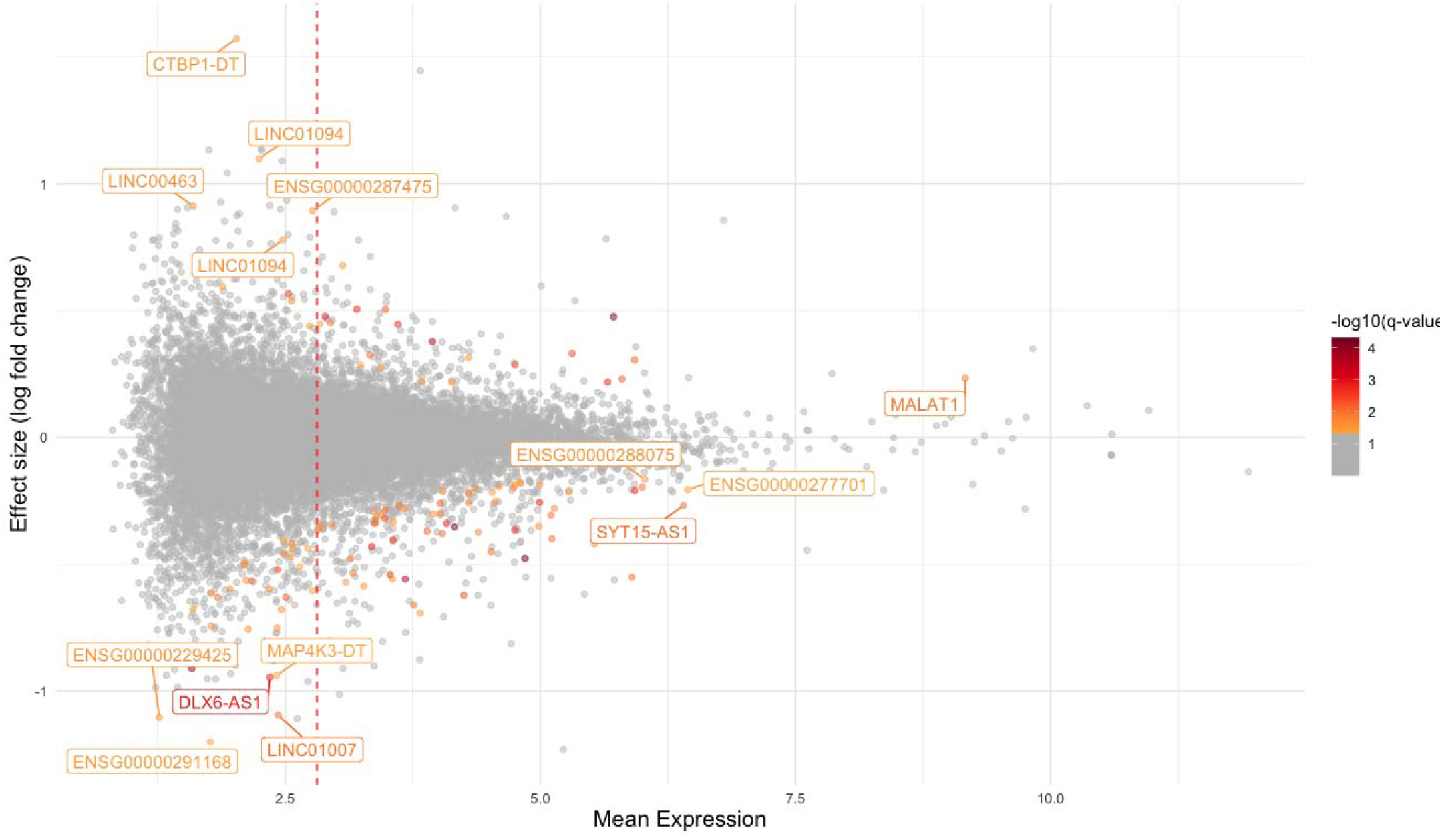
MA plot depicting the differentially expressed lncRNAs in the OPTIMA dataset. The top identified lncRNAs with significant adjusted p, higher mean expression, and effect sizes are annotated. The perpendicular red dotted line represents the average of the mean expression. The color intensity of each dot corresponds to the −log10(q) as illustrated by the color gradient legend in the figure.

The strongest effect in either direction was observed for the top ten transcripts from unique lncRNA genes, including, *CTBP1-DT* (q = 3.13E-02), *LINC01094* (q = 3.30E-02), *LINC00463* (q = 2.86E-02), *ENSG00000287475* (q = 3.13E-02), *LINC01338* (q = 3.84E-02), *ENSG00000291168* (q = 3.28E-02), *Lnc-NRIP1-2* (q = 2.88E-02), *LINC01007* (q = 1.09E-02), *DLX6-AS1* (q = 2.86E-02), and *MAP4K3-DT* (q = 2.40E-03) (**Supplementary Table S2B**). Of the 17 lncRNAs showing particularly consistent effect sizes in human brain samples according to previous work (see introduction), we found all but *BACE1-AS* to show at least nominally significant differential expression in the OPTIMA dataset (**Supplementary Table S2A**).

### Sensitivity analyses confirm that DGE results are stable across a range of methods

To assess whether and to which degree our DGE results depend on the choice of tools and parameters, we repeated the main analyses using alternative approaches. First, we quantified lncRNAs using a combination of STAR^43^ and featureCounts^44^ (instead of Kallisto, see Methods) and then compared lncRNA levels by correlation analysis. The strong Pearson’s correlation coefficients between both quantification methods in ten randomly selected controls (r = 0.95) and ten AD cases (r = 0.94) suggest that estimated lncRNA levels are stable regardless of which method was applied (**Supplementary Figure S1a-S1b)**. Second, we repeated DGE analyses using DESeq2^45^ (instead of Sleuth, see Methods), which also produced comparable results, i.e., Pearson’s correlations showed very similar mean expression levels (r = 0.93) and DGE effect sizes (r = 0.9) across the two methods (**Supplementary Figures S2a-S2b** ). Overall, these sensitivity analyses suggest that our results remain stable regardless of which approach is chosen for lncRNA and mRNA quantification or for computing DGE statistics.

### DGE analyses in other brain regions confirm differential expression for at least 58 lncRNAs

To assess the differential expression of the 121 FDR-significant lncRNAs from the OPTIMA study in other brain regions, we performed equivalent DGE analyses in data from the dorsolateral prefrontal cortex (DLPFC; ROSMAP; n = 769) and fusiform gyrus (FG; BANNER; n = 274) and compared the results from all three datasets. In ROSMAP, we identified a total of 1,431 lncRNAs to show at least nominal evidence for DGE (λ = 1.05); of these, 146 were significant after FDR control (FDR < 0.05; **Supplementary Figure S3a; Supplementary Table S3** ). In BANNER, there were 839 lncRNAs showing at least nominally significant DGE (λ = 1.07), of which 11 survived multiple testing corrections by FDR (**Supplementary Figure S3b; Supplementary Table S4)** . Comparing the ROSMAP- and BANNER-derived DGE results with the 121 FDR-significant lncRNAs from the discovery dataset (OPTIMA) revealed 58 lncRNAs (47.9%) showing at least nominally significant differential expression in the same direction in at least one of the two other brain regions anazlyzed (**Supplementary Figure S4; Supplementary Table 2B** ). This suggests that a sizeable fraction of lncRNAs is dysregulated in more than one brain region in AD cases vs. controls.

One notable difference across datasets is the number of lncRNAs to show FDR-significant DGE, i.e., OPTIMA (n = 121) and ROSMAP (n = 146) vs. BANNER (n = 11). These differences may be attributed to at least two factors: 1) the different brain regions analyzed may genuinely show a different pattern of lncRNA expression, and/or 2) the observed differences in DGE numbers reflect differences in statistical power, which in turn is correlated with sample size and RNA-seq coverage. With respect to the latter, the OPTIMA and ROSMAP data showed the highest values (mean estimated read counts: 1,532,401 and 1,390,565, respectively) while for BANNER, the coverage was only ∼50% of these values (771,200). We note that biological interpretations of our findings are only based on those lncRNAs showing consistent evidence for differential expression in meta-analyses across all three datasets (see below), minimizing potential biases created by these and possibly other factors.

### Meta-analysis reveals 356 lncRNAs showing consistent DGE across different brain regions

To increase the power and specificity of our overall approach, we next combined the DGE results obtained in the OPTIMA samples with equivalent DGE data computed for the ROSMAP and BANNER datasets by meta-analysis. Overall, this cross-cortex analysis included n=1,220 brain samples and resulted in a total of 495 significantly (FDR < 5%) differentially expressed lncRNAs in AD vs. controls. Of these, 356 lncRNAs (n[downregulated] = 225, n[upregulated] = 131) showed the same direction of effect in all three datasets **(Supplementary Table S5, Figure 3)**. As expected, the vast majority (i.e., 43/58) of the lncRNAs showing consistent DGE results across individual brain regions (see above) are also FDR-significant in the meta-analyses. The same is true for all but one (*BACE1-AS*) of the 17 previously reported brain-relevant lncRNAs (see introduction). Based on the strength and consistency of the meta-analysis results, we consider these 356 lncRNAs to represent the most compelling AD-associated lncRNAs of our study. Accordingly, we used them for downstream functional characterization analyses (see below).

**Figure 3:**
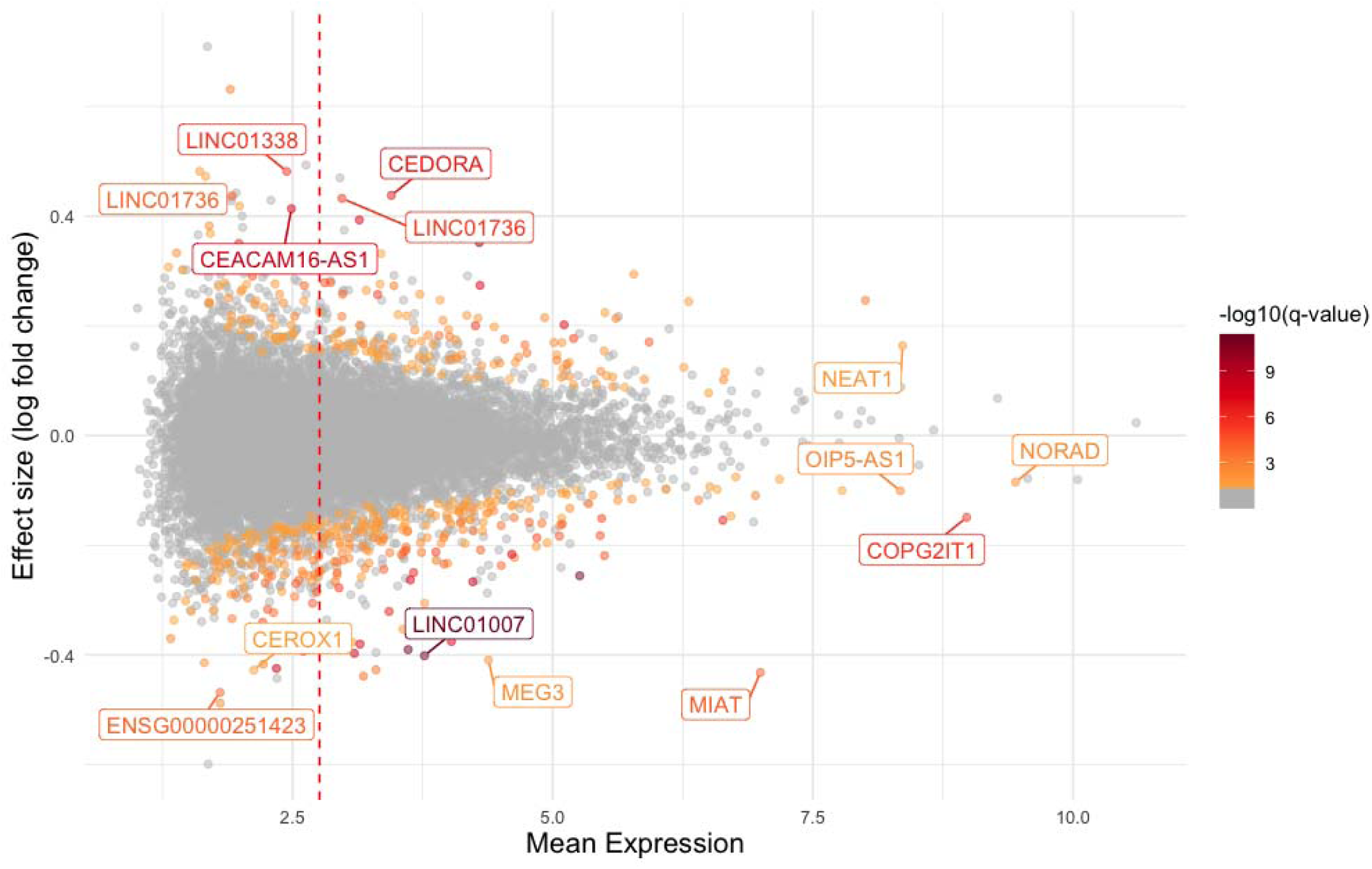
MA plot depicting the differentially expressed lncRNAs in the meta-analysis. The top identified lncRNAs present in all three meta-analyzed datasets, with significantly adjusted q and a similar direction of effect are annotated. The perpendicular red dotted line represents the average of the weighted-mean expression in all three datasets. The color intensity of each dot corresponds to the −log10(q), as illustrated by the color gradient legend in the figure.

The most significant association was observed for the top ten unique transcripts from lncRNA genes, including, *Lnc-TNFRSF13B-2* (q = 4.00E-12), *TMEM132E-DT* (q = 4.00E-12), *LINC01007* (q = 4.28E-12), *LINC00499* (q = 1.53E-11), *ENSG00000287689* (q = 2.14E-09), *Lnc-TRMT44-1* (q = 8.92E-09), *CEACAM16-AS1* (q = 1.74E-08), *CEDORA* (q = 4.24E-08), *Lnc-EFNA2-1* (q = 4.31E-08), and *THCAT155* (q = 6.25E-08).

Among the FDR-significant DGE meta-analysis results, eight had already been highlighted in previous work, including *MIAT*^35, 36^*, MAP4K3-DT* ^31^, *LY86-AS1* ^34^*, LINC01007* ^34^*, HAR1A* ^31^, *MEG3*^37^, *NEAT1* ^46^, and *LINC01094*^34^ **(Supplementary Table S5).** Other previously reported lncRNAs were either present in only two (of three) datasets [*XIST* (q = 4.09E-06);^47, 48^, *MEG8* (q = 4.44E-02);^31^, *NECTIN3-AS1*^31^ (q = 7.08E-03), and *MALAT1* (q = 4.01E-02);^49^] or only showed nominal (unadjusted for multiple comparisons) significance [*MIR7-3HG* (p = 6.08E-03);^34^, *PCA3* (p = 4.7E-02);^31^, *STARD4-AS1* (p = 4.2E-02);^31^, and *MEG9* (p = 3.45E-03);^8^] in the meta-analysis. Notably, all of these lncRNAs also showed at least nominal evidence for DGE in the OPTIMA dataset alone (see above and **Supplementary Table S2A**).

### LncRNA-mRNA correlations reveal 403 mRNAs potentially impacted by dysregulated lncRNAs

To perform a first-wave assessment of the pathogenic mechanisms potentially affected by the lncRNAs identified to show DGE, we first assigned co-expressed (and at least nominally significant DGE, see Methods) mRNAs to all 356 cross-cortex lncRNAs by pairwise correlation analyses in the entorhinal cortex. These mRNAs were then used as input into GO enrichment analyses to decipher potential functional pathways (next section). The correlation analyses identified a total of 682 lncRNA-mRNA pairs, where 58 unique lncRNAs (out of 356 lncRNAs that showed the same direction of effect in all three datasets in the meta-analysis) showed strong positive or negative correlations with 403 unique mRNAs (**Supplementary Figure S5**). Out of the 682 lncRNA-mRNA pairs (**Supplementary Table S6** ), 38 were located in *cis*, while 644 were in *trans*. The comparatively strong correlation coefficients (|0.85|) used to define these lncRNA-mRNA pairs were chosen to increase the likelihood of a direct (and potentially functionally relevant) relationship between the two RNA types. Whether or not these relationships, indeed, exist *in vivo* needs to be assessed by dedicated functional genetic experiments, which were beyond the scope of this study.

### GO enrichment analyses highlight several molecular pathways of potential relevance to AD

Next, we used the mRNAs co-expressed with top lncRNAs in GO enrichment analyses to delineate biological processes possibly affected by the most consistently differentially expressed lncRNAs. These analyses led to the identification of 28 GO terms significantly (FDR < 0.05) enriched among co-expressed mRNAs (**Supplementary Table S7** ). Many (i.e., 11 of 28) of the enriched GO annotations suggest involvements in processes related to neurogenesis (e.g., neuron development, neuron differentiation, neuron projection morphogenesis, neurogenesis, and generation of neurons), synapse function (synapse organization, anterograde trans-synaptic signaling, chemical synaptic transmission, and synapse assembly), axon development (e.g., axon development, and axonogenesis). To assess the stability of the GO enrichments, we re-computed the analyses with a different set of background genes based on different parameters (Methods). These alternative analyses resulted in largely similar GO annotation terms (**Supplementary Figures S6a-S6b** ), emphasizing the stability of our primary GO enrichment results. Together, this first pass *in silico* functional assessment appears highly plausible with respect to AD pathogenesis and opens avenues for potential future experiments.

## Discussion

This study used RNA-seq data from 1,220 carefully ascertained human brain samples to identify lncRNAs differentially expressed in AD across three different brain regions. Transcriptome-wide DGE analysis in entorhinal cortext (OPTIMA) resulted in a total of 121 lncRNAs with significantly differential expression in AD cases. To increase power, we next conducted a transcriptome-wide meta-analysis combining DGE results from entorhinal cortex (OPTIMA), dorsolateral prefrontal cortex (ROSMAP), and fusiform gyrus (BANNER), which resulted in a total of 356 FDR-significant differentially expressed lncRNAs showing the same direction of effect in all three datasets. The meta-analysis not only replicated well-established, widely reported AD-associated lncRNAs (incl. *HAR1A, LINC01094, LY86-AS1, LINC01007, MIAT, MIR7-3HG, MALAT1* , *MAP4K3-DT, MEG3, MEG8, MEG9, NEAT1, NECTIN3-AS1, STARD4-AS1, PCA3, and XIST*) but also identified dozens of highly significant lncRNAs not previously associated with AD. Post-DGE assessments using lncRNA-mRNA co-expression and gene ontology enrichment analyses revealed a number of processes of possible relevance to AD, including synaptic function, cognitive performance, axon development, and other neuron-related mechanisms. In summary, to the best of our knowledge, our comprehensive transcriptome-wide DGE study represents the largest assessment of lncRNA expression in the human brain to date. As such, it significantly extends the existing knowledge base on the topic and further emphasizes the value of considering this important and abundant class of non-coding RNA in future work.

While an exhaustive discussion of the complete list of study-wide significant lncRNAs is beyond the scope of this article, we highlight five lncRNAs not implicated in previous work showing particularly strong evidence for differential expression in our study, i.e., *Lnc-TNFRSF13B-2*, *CSRP1-AS1*, *Lnc-MIB2-1*, *Lnc-TTC5-2,* and *GSN-AS1*.

The expression of lncRNA *Lnc-TNFRSF13B-2* is highly correlated with the expression of mRNA *SMIM10L2B,* which encodes small integral membrane protein 10 like 2B (SIL2B) protein (**Supplementary Table 6**). SIL2B has been reported to be a highly abundant macrosomal protein that inhibits amyloid-beta-40 (Aβ40) fibrillation, with knockdown or cleavage of SIL2B reducing this inhibitory effect^50^. SIL2B peptides were also reported to inhibit Aβ40 aggregation in a concentration-dependent manner^50^ Overall, the overexpression of SIL2B was found to be neuroprotective against AD^50^. The second highlighted lncRNA candidate, *CSRP1-AS1,* showed a highly correlated expression with *FMNL2* (**Supplementary Table 6**). This mRNA encodes formin-like protein 2 and is interesting as it has been found to show an elevated expression in the brains of AD patients, with particular affinity to amyloid and phosphorylated tau^51^. Furthermore, its expression is particularly prominent in astroglia of AD cases with cerebrovascular pathology^51^. The third novel DGE candidate is *Lnc-MIB2-1,* whose expression was found to be highly correlated with mRNA *ZFR2* (**Supplementary Table 6**). The gene encodes (Zinc Finger RNA-Binding Protein 2), which is abundantly expressed in the brain and was suggested to be involved in dendritic branching and axon guidance^52^. The fourth highlighted lncRNA is *Lnc-TTC5-2,* which is cis-correlated with *RTN1* (**Supplementary Table 6** )*. RTN1* is primarily expressed in the nervous system, with minimal expression in peripheral tissues such as the spleen and lungs. It shares structural similarities with other RTN family members^53^. RTN1 is essential for neurofilament organization and synaptic integrity. While RTN1 deficiency alone shows no major effects, its combined loss with RTN3 causes neonatal lethality, impaired neuromuscular junctions, and axonal growth defects^54^. Previous work had already highlighted RTN1 as differentially expressed in AD vs. control brains^55^. Lastly, we highlight the DGE finding with *GSN-AS1* whose expression is highly correlated with that of *GSN* (encoding: Gelsolin), and both transcripts map to the same genomic locus (chr. 9q33.2; **Supplementary Table 6**). Previous work suggests that plasma levels of GSN are significantly elevated in AD patients but not in individuals with mild cognitive impairment (MCI)^56^. This distinction has led to the proposal that GSN could serve as a promising biomarker for developing blood-based diagnostic tests for AD^56^. Despite these promising leads for the lncRNAs highlighted above, we emphasize that the highlighted potential functional connection is based on statistical correlation only. Future work will need to assess whether or not the suspected functional link between lncRNA and mRNA expression actually does exist *in vivo*.

One noteworthy lncRNA not found to be differentially expressed in our study is *BACE1-AS*. Previous work suggests that the expression of this lncRNA is significantly higher in AD cases when compared to controls^57, 58^. As an “anti-sense” (AS) regulator of one of the key enzymes (beta-site APP cleavage protein, a.k.a. beta-secretase; BACE1) to metabolize APP into Aβ, it has been proposed to exacerbate amyloid plaque formation^25^ and even to represent a potential therapeutic target for AD. The latter notion is based on data suggesting that the knockdown of *BACE1-AS* in an animal model shows an increase in the proliferation of primary hippocampal neurons^59^. Given these consistent and functionally compelling prior findings, we explored why this lncRNA did not emerge as a top signal in our data. This revealed that *BACE1-AS* exhibited an exceedingly low expression in the brain samples analyzed here, leading to its exclusion at the QC step of our analysis. As shown in **Supplementary Figure S7a**, *BACE1-AS* expression is also low in other brain-based transcriptomics datasets, such as GTEx^60^. Notwithstanding, we explored the pattern of *BACE1-AS* expression by visualizing its read counts and – for comparison – those of *NEAT1* in four different GTEx brain regions (amygdala, cortex, hippocampus, and frontal cortex) (https://gtexportal.org/home/downloads/adult-gtex/bulk_tissue_expression) and compared them to equivalent data in OPTIMA entorhinal cortex samples (**Supplementary Figure S7b**), and found very similar expression patterns across these datasets. Comparing the expression patterns with respect to phenotype revealed that the direction of effect here (suggesting an increased expression of *BACE1-AS* in AD vs. controls; **Supplementary Figure S7c-S7d**) is in good agreement with the literature where most studies found similar patterns^22^.

The strengths of our study include, first, the utilization and combination of RNA-seq data from three comparatively large and independent datasets derived from the human brain, increasing the statistical power to detect differentially expressed lncRNAs. In this context we note that we are not aware of any prior meta-analyses combining the RNA-seq data from ROSMAP and BANNER. Second, we performed assessments across three different human brain regions, allowing us to highlight those lncRNAs showing particularly consistent effects in more than one cortical region. Third, we went to great lengths at applying conservative statistical approaches for the delineation of our primary findings. In particular, this relates to the use of PCA for an adjustment for variance in the data due to unknown technical and/or biological factors, quantification of transcriptome-wide inflation statistics, and the performance of cross-cortex meta-analyses, all with the aim of reducing the number of false-positive findings. Lastly, the overall high quality of our discovery dataset is further emphasized by the replication of many lncRNAs previously reported to be differentially expressed in AD.

Apart from these strengths, we also highlight several potential limitations. First and foremost, all analyses in this work are based on RNA-seq data generated on bulk tissue. While the interpretation of significant findings is typically not affected by this approach, it may mask cell-specific dysregulation of certain lncRNAs and, thus, overlook their link to AD pathogenesis. Second, a similar situation may have resulted from meta-analysis across different regions of the brain, resulting in masking potential region-specific lncRNA effects. However, neither of these limitations can be expected to have any bearing on the primary results of our work, as it would lead to a decrease in power. Third, RIN values in the OPTIMA dataset were comparatively low, possibly indicating a lower than usual quality of the RNA extracts. However, as we included RIN as a covariate in our statistical modeling, this should not have any major bearing on the DGE results. Fourth, while several of the suggested functional implications related to lncRNAs highlighted as differentially expressed appear highly plausible with respect to their potential role in AD pathogenesis after determining co-expression with mRNA transcripts, we note that these annotations are *in silico* in nature. Thus, to elucidate the true molecular mechanisms underlying our findings, dedicated experiments need to be conducted using applicable *in vitro* or *in vivo* models, which was beyond the scope of our study. Fifth, due to a lack of detailed clinical information, we were not able to directly include potential confounding factors such as comorbid conditions, medication history, or lifestyle factors, into our statistical models. However, as we extensively accounted for general aspects of data variability (technical, biological, or both) by principal component analysis, the impact of other unaccounted factors on the main results is likely small. Lastly, all analyzed subjects included were of Northern European (“white”) descent precluding any form of generalizability of our findings to other ancestry groups.

In conclusion, our study represents the hitherto largest – both in terms of sample size and molecular resolution – RNA-seq-based assessment of differential lncRNA expression in the AD field. By nominating 356 study-wide significant lncRNAs showing particularly consistent and robust evidence for differential expression in AD, our study substantially extends the scope and results of previous work. Future studies are needed to validate our findings and assess their relevance in AD pathogenesis by dedicated molecular experiments.

## Methods

### Description of cohorts and RNA sequencing data

#### OPTIMA

Snap-frozen post-mortem human EC slices (Brodman area BA28) were obtained from AD patients (n=95) and cognitively normal controls (n=82) from the Oxford Brain Bank in accordance with approved protocols by the Ethics Committee of the University of Oxford (ref 23/SC/0241). All participants had given prior written informed consent for the brain donation. The patients had been part of a longitudinal study of cognitive impairment called Oxford Project to Investigate Memory and Aging (OPTIMA). Detailed clinical data collection of the tissue extraction procedures^61^ and RNA-seq protocols have been described elsewhere^7^. The demographic details of the 177 OPTIMA participants used in this study are summarized in **Supplementary table S1**.

In brief, all study participants were of white European ancestry and underwent detailed physical and clinical assessments, including biannual cognitive evaluations with the Cambridge Examination of Mental Disorders of the Elderly (CAMDEX)^62^, Cambridge Cognitive Examination (CAMCOG), and Mini-Mental State Examination (MMSE). A neuropathological diagnosis of AD was based on the Consortium to Establish a Registry for Alzheimer’s Disease (CERAD) and National Institutes of Health (NIH) criteria, assessing Aβ and p-tau pathology in multiple cortical regions^63–65^. Control subjects had no clinical evidence of significant cognitive impairments along with the absence of neuropathological hallmarks suggesting the presence of AD. RNA was extracted from n = 183 OPTIMA samples and then subjected to total RNA-seq on a NovaSeq 6000 instrument (Illumina, Inc.) using 2×50bp TruSeq (Illumina, Inc.) libraries.

#### ROSMAP

RNA-seq data from dorsolateral prefrontal cortex (DLPFC) slices of n = 803 participants of the Religious Orders Study and Memory and Aging Project (ROSMAP)^40^ participants were obtained from the AD Knowledge Portal database (syn21589959, syn4164376; https://adknowledgeportal.synapse.org/). After quality control (QC), we were able to include a total of 769 **(Supplementary Table S1)** study participants (n[AD cases] = 436, n[controls] = 333] for DGE analyses. Detailed information about ROSMAP clinical procedures and RNA-seq methodology can be found elsewhere (https://doi.org/10.7303/syn3388564)^40^.

#### BANNER

Bulk RNA-seq data generated from fusiform gyrus slices of n = 289 participants of the Banner Sun Health Research Institute Brain and Body Donation Program (BANNER) were obtained from the gene expression omnibus (GEO) database using SRA study ID SRP181886 (BioProject = PRJNA516886). After QC, we were able to include a total of 274 **(Supplementary Table S1)** study participants (n[AD cases] = 206, n[controls] = 68). Detailed information about the RNA-seq methodology can be found elsewhere^41, 42^.

### LncRNA differential expression analyses

All three datasets were analyzed using the same processing and QC pipeline. First, RNA-seq reads were pseudoaligned to the human transcriptome (GRCh38.81) using Kallisto v0.50.0^66^ and normalized to transcripts per million (TPM), focusing on protein-coding (mRNA) and lncRNA isoforms using annotations from Ensembl [ensembl release-111 homo_sapiens (gtf)] (**Figure1**). Differential gene expression (DGE) analyses were conducted separately on lncRNAs as well as the combined dataset of lncRNA and mRNA mappings using the R (version 4.3.3)^67^ package Sleuth (0.30.1)^68^. To focus on the most robust portion of the transcriptome, we only retained transcripts with a minimum of 5 reads and expressed in at least 47% of the samples for subsequent analyses utilizing Sleuth’s default built-in filtration criteria. DGE analyses were performed in Sleuth and are based on a Wald test using AD status as a predictor and adjusting for potential confounders, including age at death, sex, RNA integrity number (RIN), and the first twenty principal components (PC1 to PC20, see below). For ROSMAP data, the Wald test was performed adjusting for age, sex, RIN, dementia status, and the first nine PCs (PC1 to PC9). For BANNER data, RIN values were not available. Hence, the Wald test was only adjusted for age, sex, dementia status, and the first fifteen PCs (PC1 to PC15).

Principal component analysis (PCA) was carried out across all normalized expression data to account for batch effects and other sources of technical variability not explained by the other covariates. We applied the Benjamini-Hochberg False Discovery Rate (FDR) procedure^69^ for multiple testing corrections. Study-wide significance was assumed for DGE results at FDR-corrected p-values (q-values) <5%.

### Validation of lncRNA quantification and differential expression analyses

To assess and minimize the chances of bias or otherwise skewed results possibly introduced by the choice of software used to perform read mapping and quantification as well as the downstream DGE analyses, we re-computed all steps in the primary analysis pipeline using an alternative set of tools. For this sensitivity analysis, reads were mapped to the human genome (GRCh38.81) using STAR (Version 2.7.11b)^43^ and summarized through featureCounts (Version 2.0.6)^44^. These alternative read quantifications were then compared with the primary computations obtained with Kallisto v0.50.0 ^66^. For the actual DGE analyses, quantified reads from Kallisto were alternatively analyzed using DESeq2 (1.42.1)^45^ as implemented in R (version 4.3.3)^67^. DGE results from DESeq2 (sensitivity) and Sleuth (main analysis) were then compared with each other using Pearson correlations of effect size estimates.

### Meta-analysis

DGE results from OPTIMA (discovery data) were combined with corresponding DGE results from ROSMAP and BANNER datasets by meta-analysis. The meta-analysis was performed in METAL^70^ using a fixed-effect model incorporating effect size estimates and standard errors. To reduce the inflation of the resulting test statistics, we used the genomic control option available in METAL. To account for multiple testing, we controlled the false-discovery rate (FDR; at 5%) applying the Benjamini-Hochberg procedure as implemented in R.

### LncRNA and mRNA correlation analysis

As outlined above, lncRNAs are non-coding and therefore not translated into proteins. Instead, they often influence cellular processes through transcriptional and – to a lesser extent – post-transcriptional regulation of mRNAs expression, which are the most likely mechanisms by which lncRNAs also unfold their potential effects on AD pathogenesis. To characterize the functional domains potentially affected by lncRNA-mediated dysregulation of mRNA expression, we performed systematic lncRNA-mRNA co-expression analyses for all lncRNA-mRNA pairs showing evidence for DGE in the OPTIMA dataset. Specifically, correlations were computed for lncRNAs showing study-wide significant differences (FDR <(F5%) with consistent directions of effect across all three datasets in the meta-analysis, paired with mRNAs showing at least nominally significant (p < 0.05) DGE in the OPTIMA dataset. Correlation analysis was performed using the ComplexHeatmap (2.18.0)^71^ package in R (version 4.3.3)^67^, where correlation coefficients are calculated using Pearson’s method. Downstream analysis was limited to lncRNA-mRNA pairs with high absolute correlation values (≥ 0.85 or ≤ −0.85) to ensure that only strongly co-expressed gene pairs were selected for further functional mapping.

### Gene ontology (GO) enrichment analyses

To gain further insight into the biological processes putatively affected by lncRNA-mediated dysregulated expression of mRNAs, we used highly co-expressed and correlated mRNAs for gene ontology (GO) enrichment analyses. GO enrichments were estimated utilizing the enrichGO function from clusterProfiler (4.10.1)^72^ R package (version 4.3.3)^67^. The analysis utilized brain-expressed mRNAs that showed strong correlations (absolute correlation coefficient ≥ 0.85) with brain-expressed lncRNAs as background. To assess the stability of these GO enrichments, we performed a sensitivity analysis modifying the mRNA data used as background in the GO enrichments by retaining only those brain-expressed mRNAs that showed strong correlations (absolute correlation coefficient ≥ 0.85) detected at a minimum of ten (instead of five) reads in at least 47% of the samples. This reduced set of mRNAs was then used as background for the GO enrichment analyses to assess how the change affected the results.

## Data Availability

All data produced in the present study are available upon reasonable request to the authors.

## Acknowledgements

This work was funded by the Deutsche Forschungsgemeinschaft (to L.B., grant ID# 391523883) and the Cure Alzheimer’s Fund (to L.B., as part of the “CIRCUITS-AD” project). Additional support was provided by the DFG Research Infrastructure NGS_CC (project 407495230) as part of the Next Generation Sequencing Competence Network (#423957469). NGS analyses were carried out at the Competence Centre for Genomic Analysis (Kiel). The use of OPTIMA samples was made possible via the Oxford Brain Bank, supported by Brains for Dementia Research (BDR) (Alzheimer Society and Alzheimer Research UK) and the NIHR Oxford Biomedical Research Centre. C.M.L. was supported by the Heisenberg Program of the DFG (LI 2654/4–1). We sincerely appreciate the participants of the Religious Order Study and the Memory and Aging Project (ROSMAP) for their valuable contributions. We also extend our gratitude to the ROSMAP principal investigators and dedicated staff for their efforts in generating and providing the data. Additionally, we are grateful to the participants of the Banner Sun Health Research Institute Brain and Body Donation Program (BANNER) in Sun City, Arizona. We also thank the BANNER principal investigators and staff for their commitment to producing and sharing the data.

## Author contributions

Overall study design: L.B., M.M.A., and M.S.

Data acquisition & processing: J.F., L.F., L.P., V.D., C.M.L., M.M.A. Statistical analyses: M.M.A., M.S.

Interpretation of findings: M.M.A., M.S., L.B. Writing of the first draft: M.M.A. L.B.

Review, editing, and approval of final draft: All authors.

## Materials & Correspondence

Prof. Lars Bertram

## Supplementary Materials

**Supplementary Figure S1:**
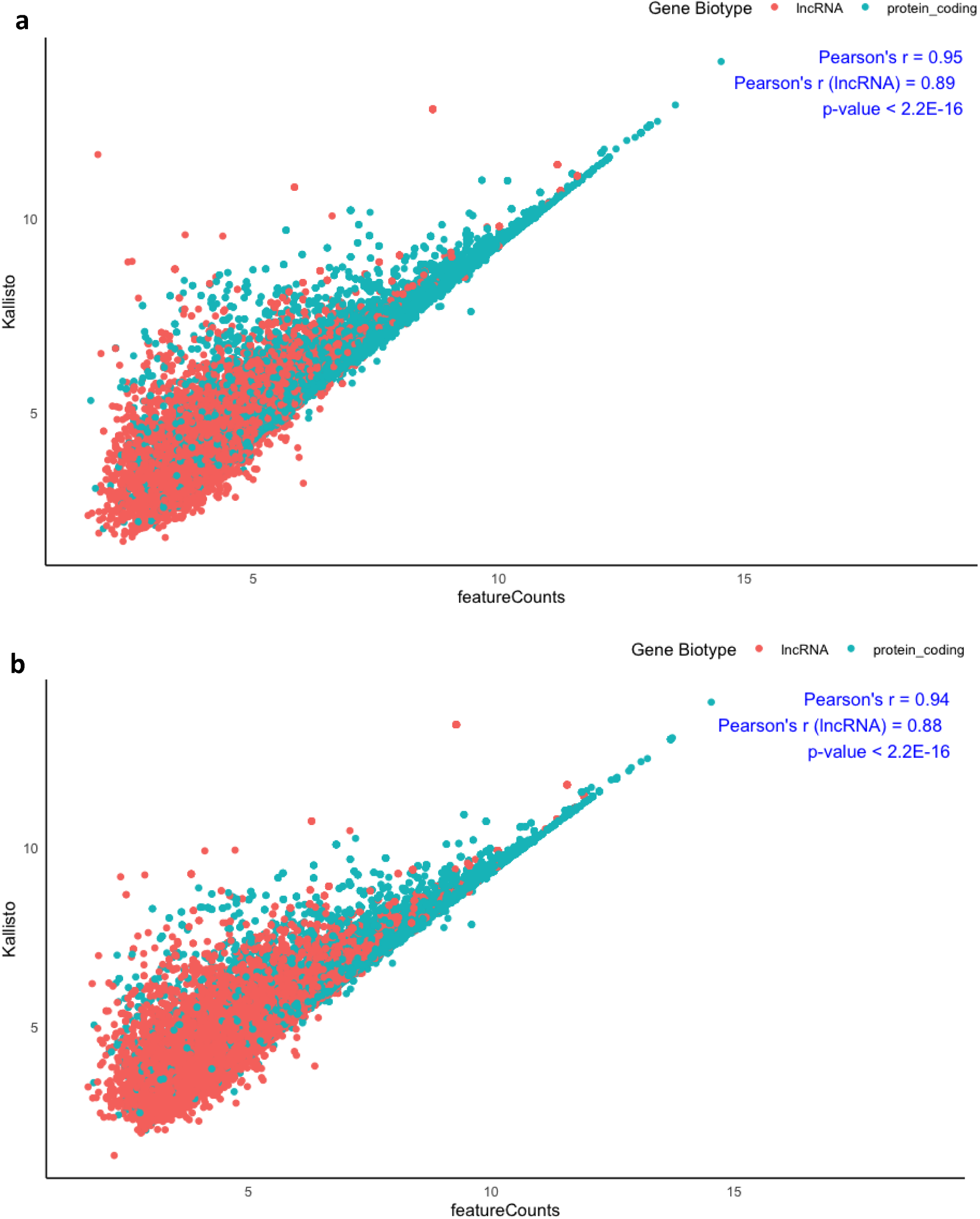
Comparison of transcript quantification done by STAR followed b y featureCounts with that obtained from Kallisto. a) Correlation of mean log quantification in tne controls. b) Correlation of mean log quantification in ten AD cases.

**Supplementary Figure S2:**
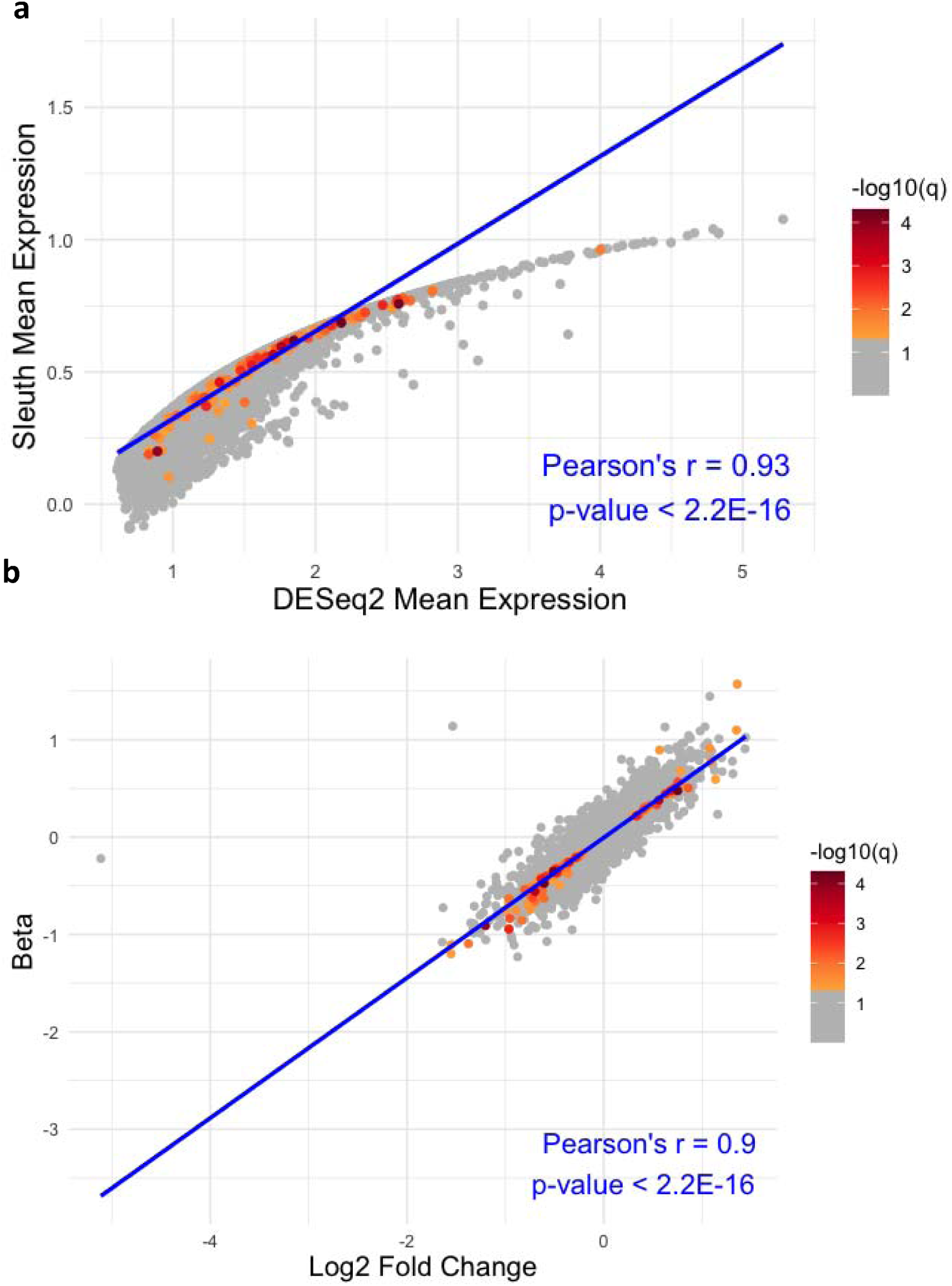
Comparison of differential gene expression (DGE) results for lncRNA using Sleuth and DESeq2. **a)** Correlation of calculated mean expression during DGE betwee n Sleuth and DESeq2. **b)** Correlation of calculated effect size during DGE between Sleuth an d DESeq2.

**Supplementary Figure S3:**
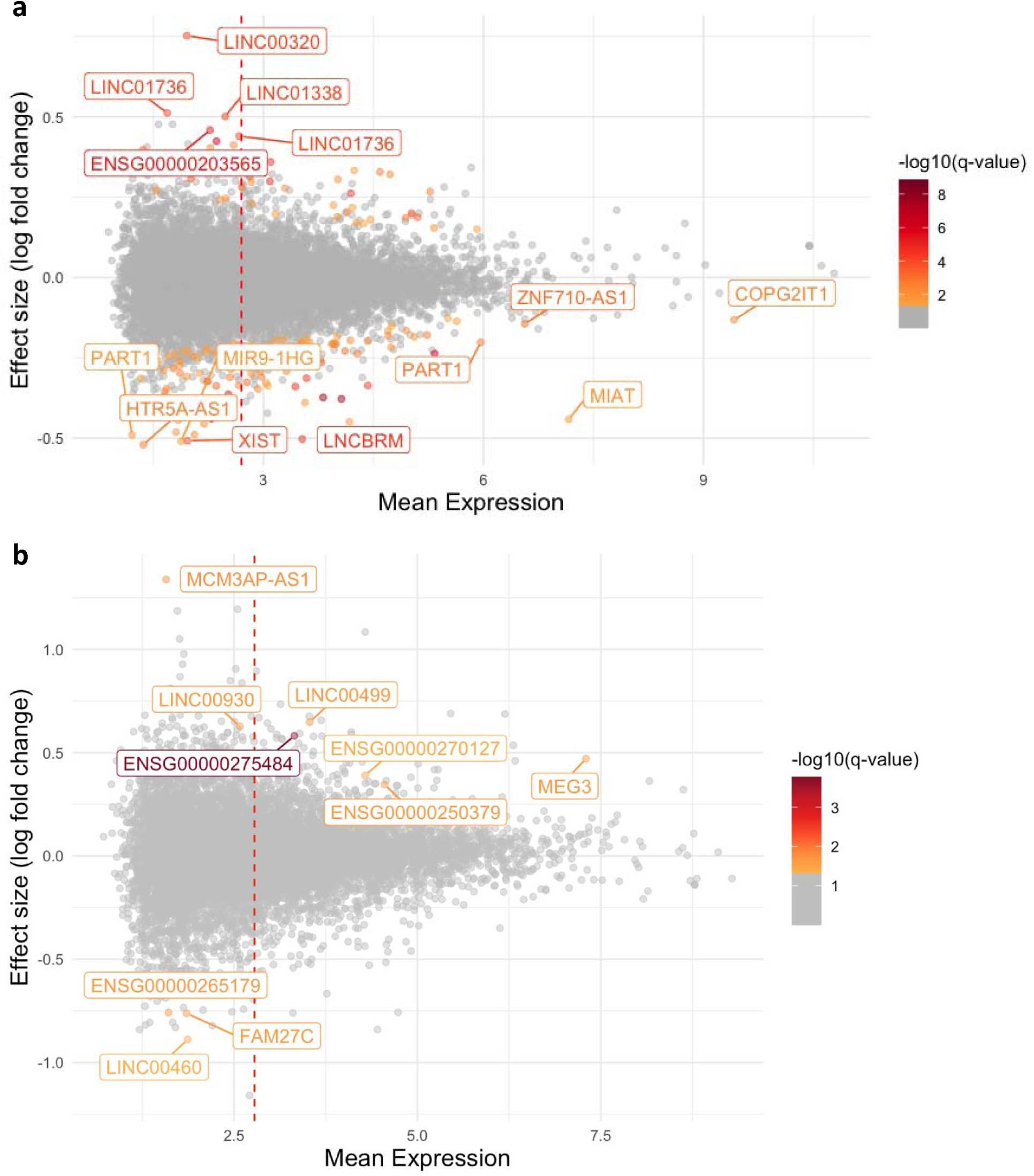
MA plot depicting the differentially expressed lncRNAs in the ROSMAP **(a)** and BANNER **(b)** datasets. The top identified lncRNAs with significant adjusted p-values, higher mean expression, and effect sizes are annotated. The perpendicular red dotted line represents the average of the mean expression. The color intensity of each dot correspond to the −log10(q-value), as illustrated by the color gradient legend in the figure.

**Supplementary Figure S4:**
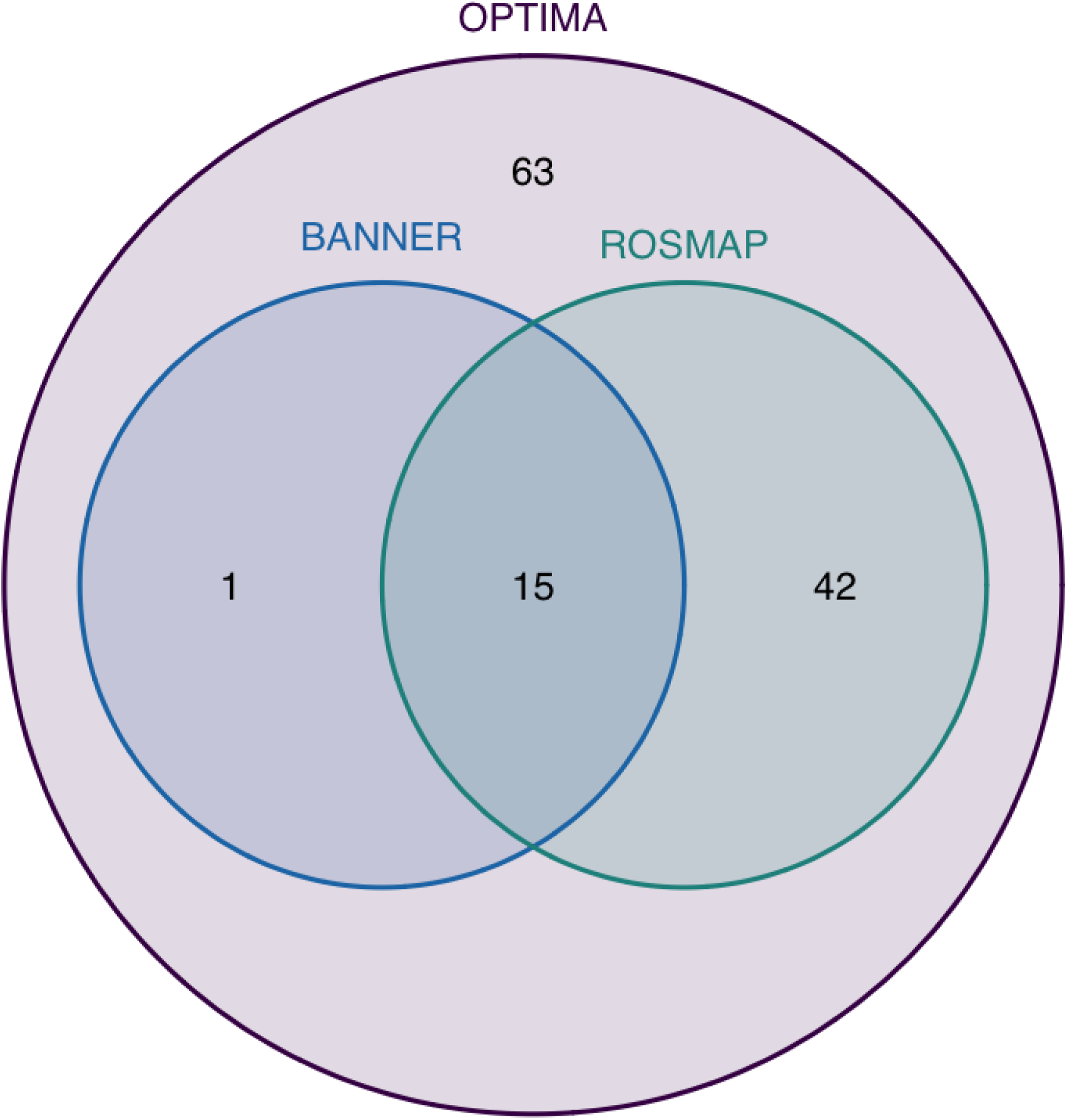
Venn diagram depicting the overlap across study-wide significan t lncRNA DGE results in OPTIMA and at least nominally significant lncRNA DGE results wit h identical effect direction in ROSMAP and/or BANNER datasets. For a full list of overlappin g lncRNAs see Supplementary Table S2B.

**Supplementary Figure S5:**
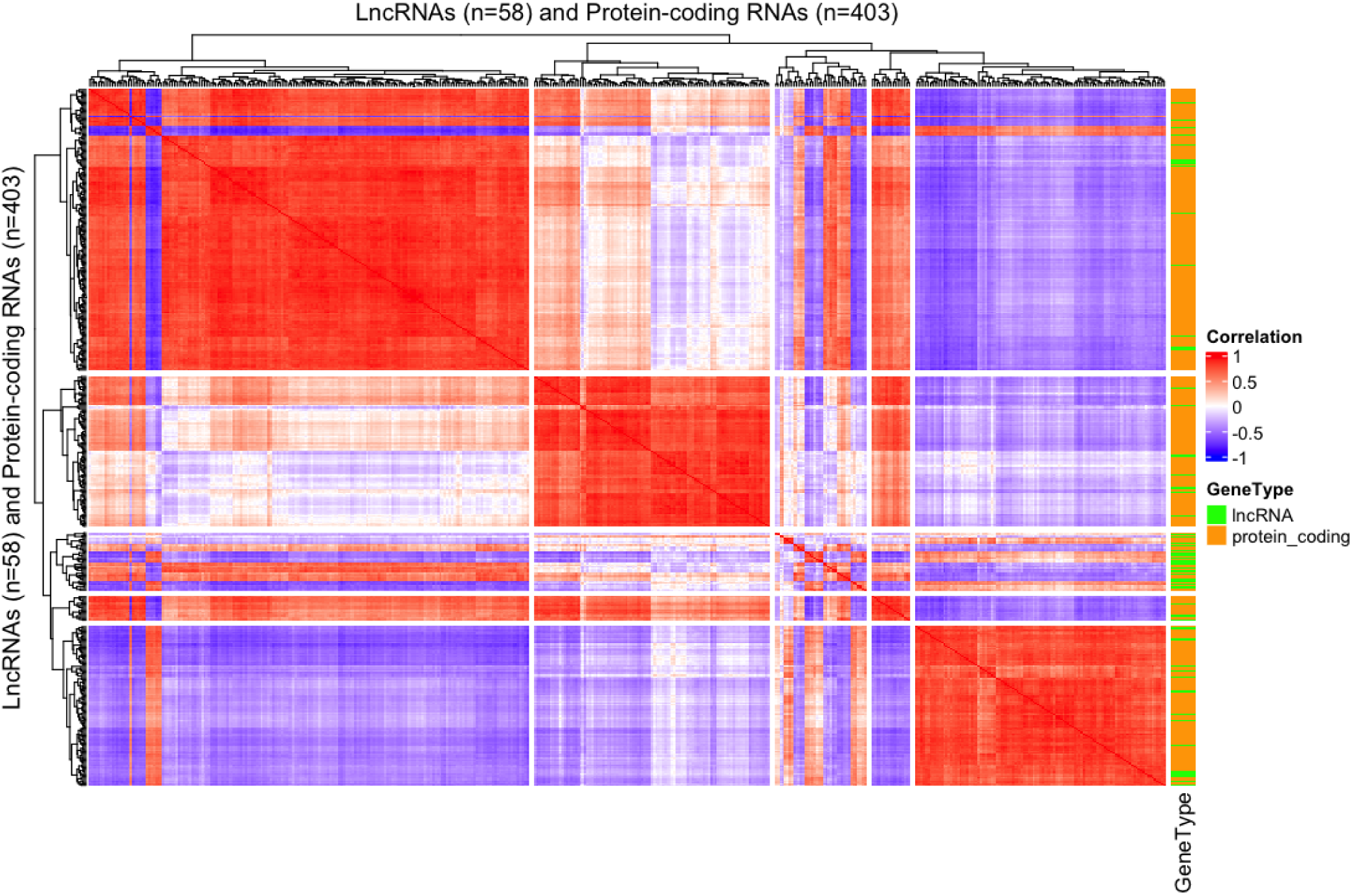
Heatmap showingstatistical correlation of lncR NAs and mRNAs pair with high absolute correlation values (≥ 0.85 or ≤ −0.85). The correlation analyses identified a total of 682 lncRNA-mRNA pairs, where 58 unique lncRNAs (out of 356 lncRNAs that showedeth same direction of effect in all three datasets in the meta-analysis) showed strong (≥ 0.85 or ≤−0.85) correlations with 403 unique mRNAs. For the list of 682 co-expressed lncRNA-mRNA pai,rs see Supplementary Table S6.

**Supplementary Figure S6:**
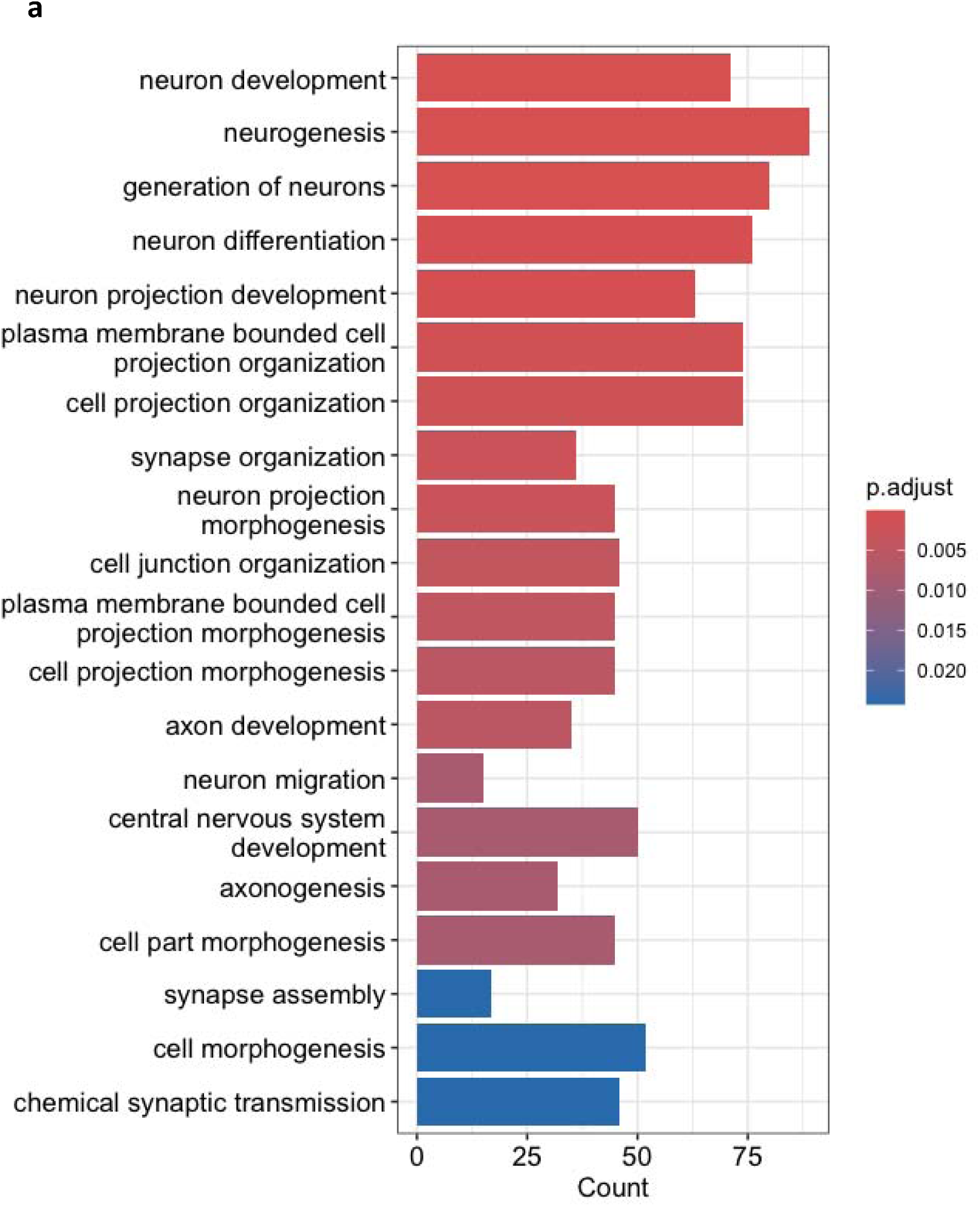
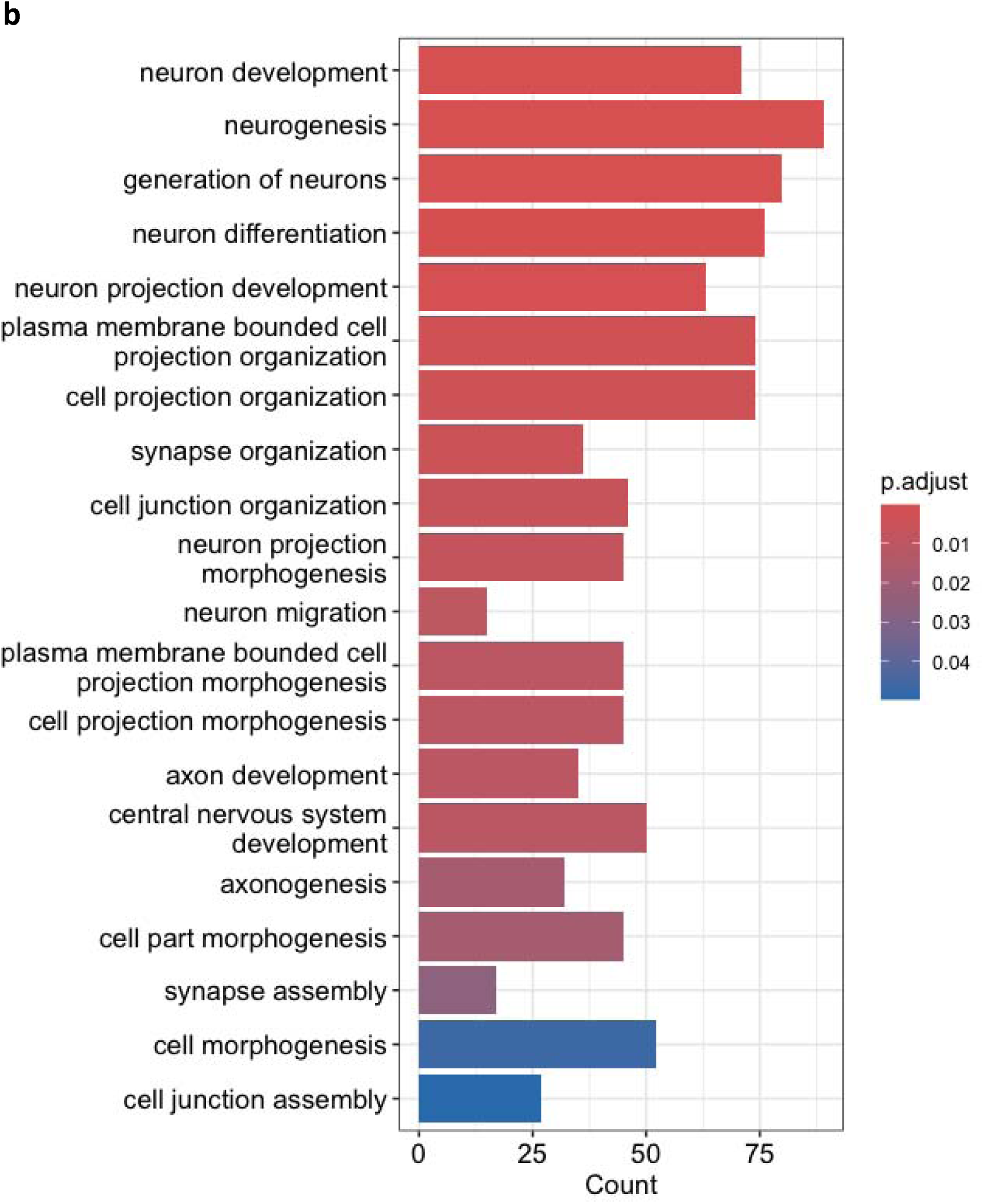
Visualization of the enrichment of biological processes through GO annotation. Panels a) and b) compare the biological processes identified following sensitivit y analysis. **a)** Visualization of the identified biological processes utilizing 403 co-expressed mRNsA against 2,990 background mRNAs for which at least five reads in 47% of the samples wer e detected (Methods). **b)** Visualization of the identified biological processes utilizing 403 co -expressed mRNAs against 2,811 background mRNAs for which at least 10 reads in 47% of th e samples were detected (Methods).

**Supplementary Figure S7:**
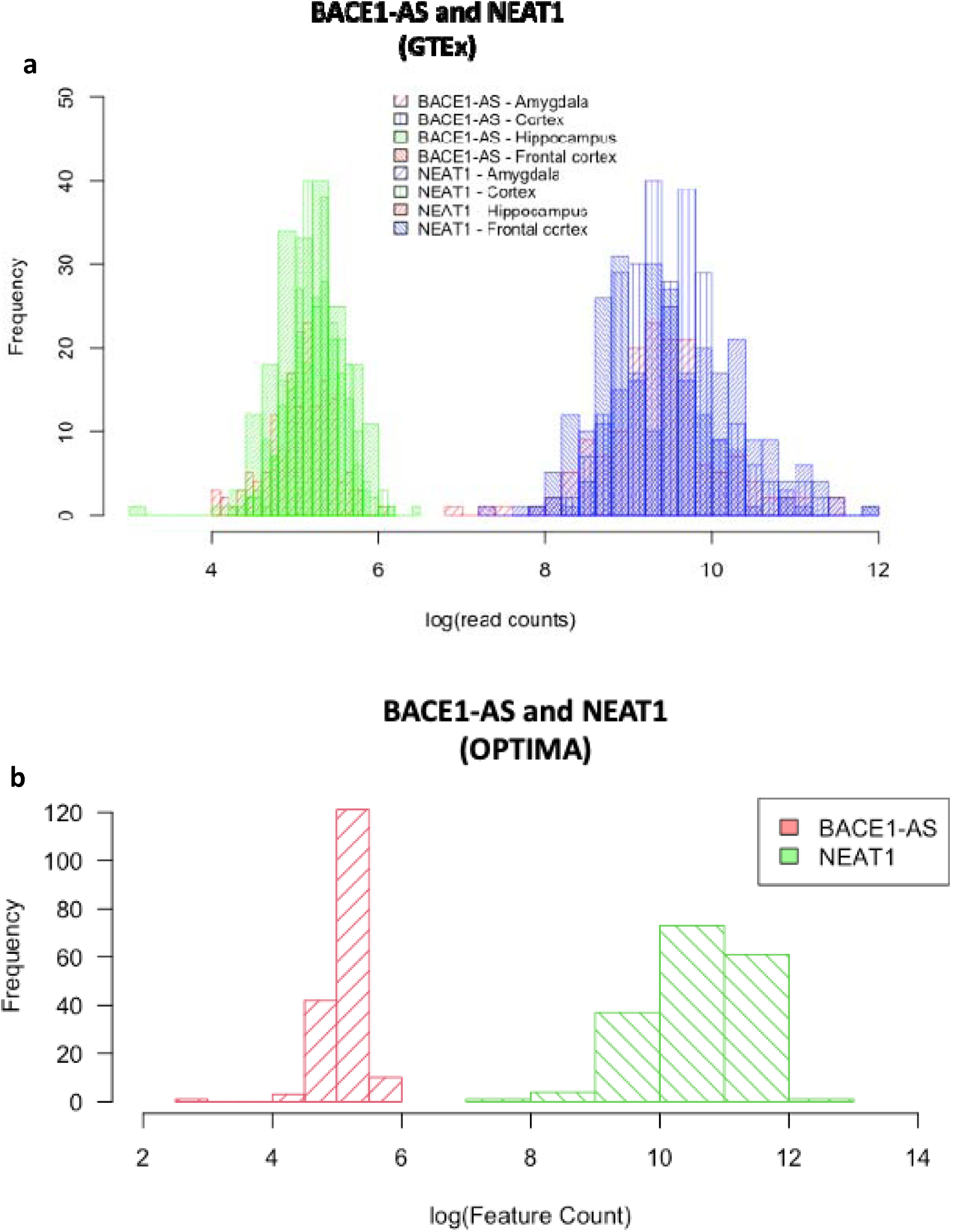
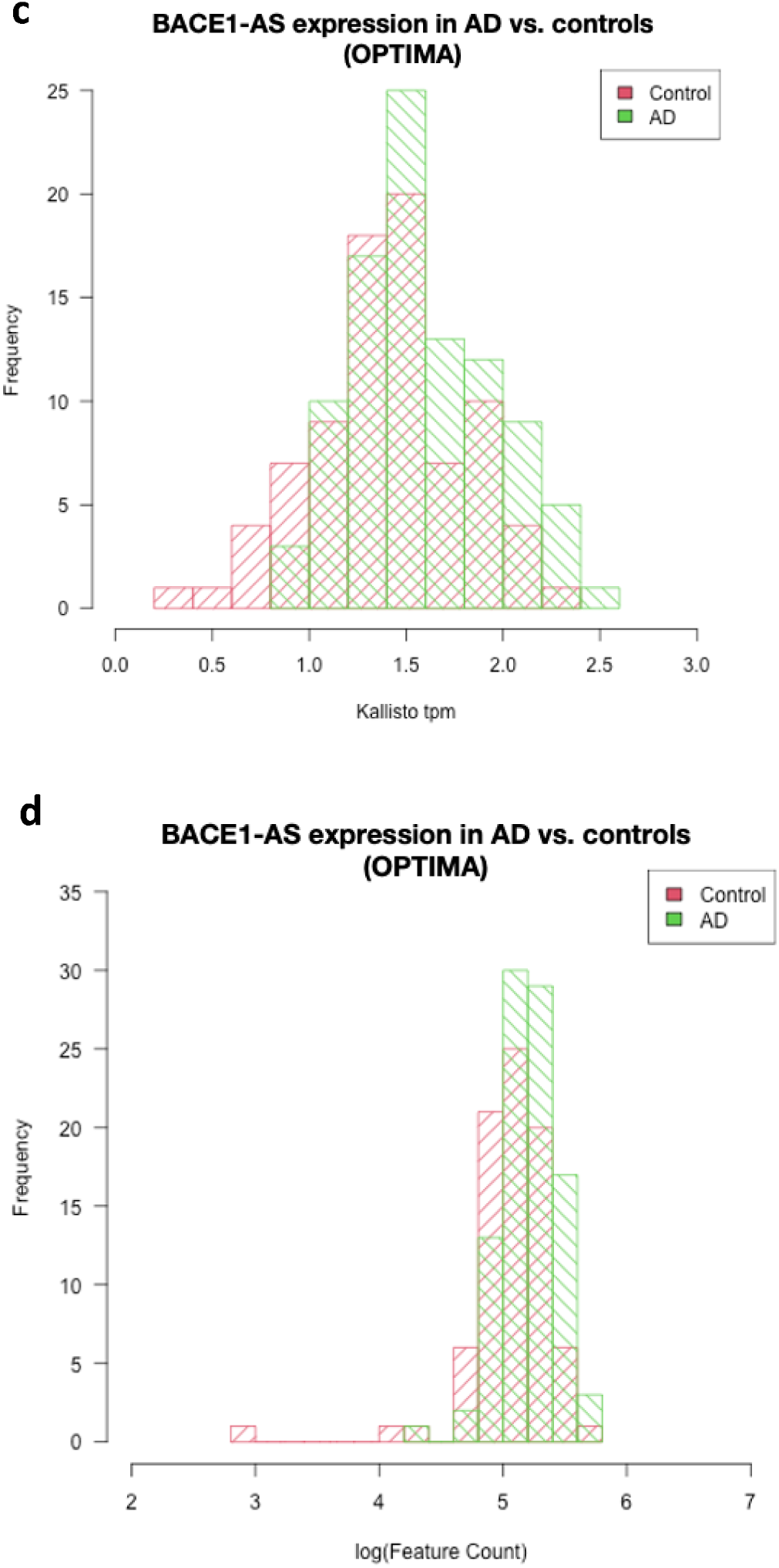
**a)** Comparison of read counts between abundantly expressed*NEAT1* and less abundantly expressed *BACE1-AS* using GTEx data from four brain regions. **b)** Comparison of read counts between *NEAT1* and *BACE1-AS1* in the entorhinal cortex (OPTIMA dataset, this study). **c)** Comparison of the *BACE1-AS1* Kallisto and **d)** featureCount rea d quantifications between AD cases and controls in the entorhinal cortex (OPTIMA dataset, thi study).

## References

1. Aslam MM, et al. Genome-wide analysis identifies novel loci influencing plasma apolipoprotein E concentration and Alzheimer’s disease risk. Molecular Psychiatry 28, 4451–4462 (2023).

2. Harper JD, et al. Genome-Wide Association Study of Incident Dementia in a Community-Based Sample of Older Subjects. J Alzheimers Dis 88, 787–798 (2022).

3. Knopman DS, et al. Alzheimer disease. Nature Reviews Disease Primers 7, 33 (2021).

4. Gorijala P, et al. Alzheimer’s polygenic risk scores are associated with cognitive phenotypes in Down syndrome. Alzheimer’s & Dementia 20, 1038–1049 (2024).

5. von Strauss E, Viitanen M, De Ronchi D, Winblad B, Fratiglioni L. Aging and the Occurrence of Dementia: Findings From a Population-Based Cohort With a Large Sample of Nonagenarians. Archives of Neurology 56, 587–592 (1999).

6. Guo L, Zhong MB, Zhang L, Zhang B, Cai D. Sex Differences in Alzheimer’s Disease: Insights From the Multiomics Landscape. Biological Psychiatry 91, 61–71 (2022).

7. Dobricic V, et al. Differential microRNA expression analyses across two brain regions in Alzheimer’s disease. Translational Psychiatry 12, 352 (2022).

8. Wang E, Lemos Duarte M, Rothman LE, Cai D, Zhang B. Non-coding RNAs in Alzheimer’s disease: perspectives from omics studies. Human Molecular Genetics 31, R54–R61 (2022).

9. Nair L, Chung H, Basu U. Regulation of long non-coding RNAs and genome dynamics by the RNA surveillance machinery. Nature Reviews Molecular Cell Biology 21, 123–136 (2020).

10. Kopp F, Mendell JT. Functional Classification and Experimental Dissection of Long Noncoding RNAs. Cell 172, 393–407 (2018).

11. Derrien T, et al. The GENCODE v7 catalog of human long noncoding RNAs: Analysis of their gene structure, evolution, and expression. Genome Research 22, 1775–1789 (2012).

12. Zuckerman B, Ulitsky I. Predictive models of subcellular localization of long RNAs. Rna 25, 557–572 (2019).

13. Brown JA, Valenstein ML, Yario TA, Tycowski KT, Steitz JA. Formation of triple-helical structures by the 31-end sequences of MALAT1 and MENβ noncoding RNAs. Proceedings of the National Academy of Sciences 109, 19202–19207 (2012).

14. Wilusz JE, JnBaptiste CK, Lu LY, Kuhn C-D, Joshua-Tor L, Sharp PA. A triple helix stabilizes the 31 ends of long noncoding RNAs that lack poly (A) tails. Genes & development 26, 2392–2407 (2012).

15. Djebali S, et al. Landscape of transcription in human cells. Nature 489, 101–108 (2012).

16. Cabili MN, et al. Localization and abundance analysis of human lncRNAs at single-cell and single-molecule resolution. Genome biology 16, 1–16 (2015).

17. Hezroni H, Koppstein D, Schwartz MG, Avrutin A, Bartel DP, Ulitsky I. Principles of long noncoding RNA evolution derived from direct comparison of transcriptomes in 17 species. Cell reports 11, 1110–1122 (2015).

18. Ulitsky I. Evolution to the rescue: using comparative genomics to understand long non-coding RNAs. Nature Reviews Genetics 17, 601–614 (2016).

19. Xiong W, Lu L, Li J. Long non-coding RNAs with essential roles in neurodegenerative disorders. Neural Regeneration Research 19, (2024).

20. Statello L, Guo C-J, Chen L-L, Huarte M. Gene regulation by long non-coding RNAs and its biological functions. Nature Reviews Molecular Cell Biology 22, 96–118 (2021).

21. Wei CW, Luo T, Zou SS, Wu AS. The Role of Long Noncoding RNAs in Central Nervous System and Neurodegenerative Diseases. Front Behav Neurosci 12, 175 (2018).

22. Shobeiri P, et al. Circulating long non-coding RNAs as novel diagnostic biomarkers for Alzheimer’s disease (AD): A systematic review and meta-analysis. PLOS ONE 18, e0281784 (2023).

23. Ren Z, Chu C, Pang Y, Cai H, Jia L. A Group of Long Non-coding RNAs in Blood Acts as a Specific Biomarker of Alzheimer’s Disease. Molecular Neurobiology 60, 566–575 (2023).

24. Lopez-Font I, Boix CP, Zetterberg H, Blennow K, Sáez-Valero J. Characterization of Cerebrospinal Fluid BACE1 Species. Molecular Neurobiology 56, 8603–8616 (2019).

25. Faghihi MA, et al. Expression of a noncoding RNA is elevated in Alzheimer’s disease and drives rapid feed-forward regulation of β-secretase. Nature medicine 14, 723–730 (2008).

26. Mus E, Hof PR, Tiedge H. Dendritic BC200 RNA in aging and in Alzheimer’s disease. Proceedings of the National Academy of Sciences 104, 10679–10684 (2007).

27. Massone S, et al. 17A, a novel non-coding RNA, regulates GABA B alternative splicing and signaling in response to inflammatory stimuli and in Alzheimer disease. Neurobiology of disease 41, 308–317 (2011).

28. Ciarlo E, et al. An intronic ncRNA-dependent regulation of SORL1 expression affecting Aβ formation is upregulated in post-mortem Alzheimer’s disease brain samples. Disease models & mechanisms 6, 424–433 (2013).

29. Massone S, et al. NDM29, a RNA polymerase III-dependent non coding RNA, promotes amyloidogenic processing of APP and amyloid β secretion. Biochimica et Biophysica Acta (BBA)-Molecular Cell Research 1823, 1170–1177 (2012).

30. Chanda K, Jana NR, Mukhopadhyay D. Long non-coding RNA MALAT1 protects against Aβ1–42 induced toxicity by regulating the expression of receptor tyrosine kinase EPHA2 via quenching miR-200a/26a/26b in Alzheimer’s disease. Life Sciences 302, 120652 (2022).

31. Filomena E, Picardi E, Tullo A, Pesole G, D’Erchia AM. Identification of deregulated lncRNAs in Alzheimer’s disease: an integrated gene co-expression network analysis of hippocampus and fusiform gyrus RNA-seq datasets. Frontiers in Aging Neuroscience 16, (2024).

32. Garcia AX, Xu J, Cheng F, Ruppin E, Schäffer AA. Altered gene expression in excitatory neurons is associated with Alzheimer’s disease and its higher incidence in women. Alzheimers Dement (N Y) 9, e12373 (2023).

33. Ng B, et al. Spatial Expression of Long Non-Coding RNAs in Human Brains of Alzheimer’s Disease. (ed(eds). Cold Spring Harbor Laboratory (2024).

34. Cao M, Li H, Zhao J, Cui J, Hu G. Identification of age- and gender-associated long noncoding RNAs in the human brain with Alzheimer’s disease. Neurobiol Aging 81, 116–126 (2019).

35. Jiang Q, et al. Long non-coding RNA-MIAT promotes neurovascular remodeling in the eye and brain. Oncotarget 7, 49688–49698 (2016).

36. Ou G-y, Lin W-w, Zhao W-j. Construction of Long Noncoding RNA-Associated ceRNA Networks Reveals Potential Biomarkers in Alzheimer’s Disease. Journal of Alzheimer’s Disease 82, 169–183 (2021).

37. Balusu S, et al. MEG3 activates necroptosis in human neuron xenografts modeling Alzheimer’s disease. Science 381, 1176–1182 (2023).

38. Amlie-Wolf A, et al. Inferring the Molecular Mechanisms of Noncoding Alzheimer’s Disease-Associated Genetic Variants. J Alzheimers Dis 72, 301–318 (2019).

39. Prokopenko D, et al. Whole-genome sequencing reveals new Alzheimer’s disease-associated rare variants in loci related to synaptic function and neuronal development. Alzheimers Dement 17, 1509–1527 (2021).

40. Bennett DA, Buchman AS, Boyle PA, Barnes LL, Wilson RS, Schneider JA. Religious Orders Study and Rush Memory and Aging Project. J Alzheimers Dis 64, S161–S189 (2018).

41. Beach TG, et al. Arizona Study of Aging and NeurodegenerativeDisorders and Brain and Body Donation Program. Neuropathology 35, 354–389 (2015).

42. Srinivasan K, et al. Alzheimer’s Patient Microglia Exhibit Enhanced Aging and Unique Transcriptional Activation. Cell Reports 31, 107843 (2020).

43. Dobin A, et al. STAR: ultrafast universal RNA-seq aligner. Bioinformatics 29, 15–21 (2013).

44. Liao Y, Smyth GK, Shi W. featureCounts: an efficient general purpose program for assigning sequence reads to genomic features. Bioinformatics 30, 923–930 (2014).

45. Love MI, Huber W, Anders S. Moderated estimation of fold change and dispersion for RNA-seq data with DESeq2. Genome Biology 15, 550 (2014).

46. Khodayi M, Khalaj-Kondori M, Hoseinpour Feizi MA, Jabarpour Bonyadi M, Talebi M. Plasma lncRNA profiling identified BC200 and NEAT1 lncRNAs as potential blood-based biomarkers for late-onset Alzheimer’s disease. Excli j 21, 772–785 (2022).

47. Huang B, Ou G-y, Zhang N. Identification of key regulatory molecules in the early development stage of Alzheimer’s disease. Journal of Cellular and Molecular Medicine 28, e18151 (2024).

48. Chanda K, Mukhopadhyay D. LncRNA Xist, X-chromosome Instability and Alzheimer’s Disease. Curr Alzheimer Res 17, 499–507 (2020).

49. Zhuang J, et al. Long noncoding RNA MALAT1 and its target microRNA-125b are potential biomarkers for Alzheimer’s disease management via interactions with FOXQ1, PTGS2 and CDK5. Am J Transl Res 12, 5940–5954 (2020).

50. Wang C, et al. Secreted endogenous macrosomes reduce Aβ burden and ameliorate Alzheimer’s disease. Science Advances 9, (2023).

51. Lee AJ, et al. FMNL2 regulates gliovascular interactions and is associated with vascular risk factors and cerebrovascular pathology in Alzheimer’s disease. Acta Neuropathol 144, 59–79 (2022).

52. Manoharan A, Ballambattu VB, Palani R. Genetic architecture of preeclampsia. Clinica Chimica Acta 558, 119656 (2024).

53. Shi Q, Ge Y, He W, Hu X, Yan R. RTN1 and RTN3 protein are differentially associated with senile plaques in Alzheimer’s brains. Scientific Reports 7, 6145 (2017).

54. Zhou J, et al. Reticulons 1 and 3 are essential for axonal growth and synaptic maintenance associated with intellectual development. Human Molecular Genetics 32, 2587–2599 (2023).

55. Liao W, et al. Identification of candidate genes associated with clinical onset of Alzheimer’s disease. Front Neurosci 16, 1060111 (2022).

56. Wang Y, et al. A blood-based composite panel that screens Alzheimer’s disease. Biomarker Research 11, 53 (2023).

57. Fotuhi SN, Khalaj-Kondori M, Hoseinpour Feizi MA, Talebi M. Long Non-coding RNA BACE1-AS May Serve as an Alzheimer’s Disease Blood-Based Biomarker. Journal of Molecular Neuroscience 69, 351–359 (2019).

58. Faghihi MA, et al. Expression of a noncoding RNA is elevated in Alzheimer’s disease and drives rapid feed-forward regulation of beta-secretase. Nat Med 14, 723–730 (2008).

59. Zhang W, Zhao H, Wu Q, Xu W, Xia M. Knockdown of BACE1-AS by siRNA improves memory and learning behaviors in Alzheimer’s disease animal model. Exp Ther Med 16, 2080–2086 (2018).

60. Lonsdale J, et al. The Genotype-Tissue Expression (GTEx) project. Nature Genetics 45, 580–585 (2013).

61. Clarke R, Smith AD, Jobst KA, Refsum H, Sutton L, Ueland PM. Folate, vitamin B12, and serum total homocysteine levels in confirmed Alzheimer disease. Arch Neurol 55, 1449–1455 (1998).

62. Roth M, et al. CAMDEX. A standardised instrument for the diagnosis of mental disorder in the elderly with special reference to the early detection of dementia. Br J Psychiatry 149, 698–709 (1986).

63. Braak H, Braak E. Neuropathological stageing of Alzheimer-related changes. Acta Neuropathologica 82, 239–259 (1991).

64. Montine TJ, et al. National Institute on Aging-Alzheimer’s Association guidelines for the neuropathologic assessment of Alzheimer’s disease: a practical approach. Acta Neuropathol 123, 1–11 (2012).

65. Mirra SS, et al. The Consortium to Establish a Registry for Alzheimer’s Disease (CERAD). Part II. Standardization of the neuropathologic assessment of Alzheimer’s disease. Neurology 41, 479–486 (1991).

66. Bray NL, Pimentel H, Melsted P, Pachter L. Near-optimal probabilistic RNA-seq quantification. Nature Biotechnology 34, 525–527 (2016).

67. Team RC. R: A language and environment for statistical computing. R Foundation for Statistical Computing. (ed(eds) (2024).

68. Pimentel H, Bray NL, Puente S, Melsted P, Pachter L. Differential analysis of RNA-seq incorporating quantification uncertainty. Nat Methods 14, 687–690 (2017).

69. Benjamini Y, Hochberg Y. Controlling the False Discovery Rate: A Practical and Powerful Approach to Multiple Testing. Journal of the Royal Statistical Society: Series B (Methodological) 57, 289–300 (1995).

70. Willer CJ, Li Y, Abecasis GR. METAL: fast and efficient meta-analysis of genomewide association scans. Bioinformatics 26, 2190–2191 (2010).

71. Gu Z, Eils R, Schlesner M. Complex heatmaps reveal patterns and correlations in multidimensional genomic data. Bioinformatics 32, 2847–2849 (2016).

72. Wu T, et al. clusterProfiler 4.0: A universal enrichment tool for interpreting omics data. The Innovation 2, 100141 (2021).

